# CSF complement proteins are elevated in prodromal to moderate AD patients and are not altered by the anti-tau antibody semorinemab

**DOI:** 10.1101/2024.04.13.24305768

**Authors:** Cosme Sandoval, Julie Lee, Balazs Toth, Rajini Nagaraj, Stephen Schauer, Jennifer Hoffman, Gwendlyn Kollmorgen, Cecilia Monteiro, Edmond Teng, Jesse E Hanson, Felix L Yeh, Johnny Gutierrez, Anne Biever

**Author notes:** equal contribution (Cosme Sandoval, Julie Lee).

## Abstract

**INTRODUCTION:** Growing evidence suggests a role for neuroinflammation in Alzheimer’s Disease (AD) pathogenesis. We investigated the complement system, a component of innate immunity, in AD patient cerebrospinal fluid (CSF) and evaluated its modulation by the anti-tau antibody semorinemab.

**METHODS:** Immunoassays were applied to measure intact (inactive) and cleaved (active) CSF complement proteins C4, Factor B and C3 in AD patients and a separate cognitively normal (CN) cohort.

**RESULTS:** All measured CSF complement proteins were increased in AD vs CN subjects, with cleaved C4 (C4a) displaying the most robust increase. Finally, semorinemab did not have a significant pharmacodynamic effect on CSF complement proteins.

**DISCUSSION:** Elevated levels of CSF intact/cleaved C4 and C3 are indicative of classical complement pathway activation in AD. Despite showing a reduction in CSF soluble tau species, semorinemab did not impact complement protein levels or activity. Further studies are needed to determine the value of complement proteins as neuroinflammation biomarkers in AD.

## 1. INTRODUCTION

Alzheimer’s Disease (AD), the predominant form of dementia, is neuropathologically characterized by extracellular amyloid β (Aβ) plaques and intracellular neurofibrillary tau tangles (NFTs) [1]. In addition to these main pathological hallmarks, activated glial cells have been observed around affected neurons and Aβ plaques [2]. Recent genome-wide association studies (GWAS) have identified AD risk genes that are enriched in glia and play a role in immune responses [2]. These findings suggest that neuroinflammation may represent a third hallmark of AD pathogenesis [3,4].

The complement system is a key component of the innate immune system and comprises three pathways. While the classical pathway is activated by immune complexes and other structures (e.g., neuronal synapses), lectin pathway activation is promoted by carbohydrates on pathogen surfaces. Both separate pathways induce the cleavage of C4 into C4a and C4b, followed by the cleavage of C2 and the formation of a C3 convertase, cleaving C3 into C3a and C3b. The alternative pathway involves interactions between spontaneously hydrolyzed C3 with Factor B and Factor D, leading to the formation of a C3 convertase. All pathways converge onto the formation of a C5 convertase, cleaving C5 into C5a and C5b and initiating the osmolysis of cells or pathogens. C3a and C5a act as pro-inflammatory anaphylatoxins, while the opsonins C4b and C3b promote the phagocytosis of invading organisms and altered self [5].

Transcriptomics and proteomics analyses reveal increased brain expression of classical complement pathway components in humans with AD and mouse models of amyloidosis and tauopathy [6,7]. The inhibition/deletion of complement proteins C1Q and C3 rescues synapse loss in amyloidosis and tauopathy mouse models, suggesting that the classical complement pathway contributes to glia-mediated synapse phagocytosis in AD [7–9]. There is thus a growing interest to further characterize complement activation and evaluate complement-targeting therapeutics in AD. Previous publications investigating complement proteins in AD cerebrospinal fluid (CSF) showed mixed results, with some, though not all, studies reporting increased CSF complement levels in AD patients (vs. healthy controls) [10,11]. Potential drivers of these inconsistencies include small study sizes and differences in diagnostic criteria, preanalytical procedures, and analytical methods. Notably, only a limited number of studies investigated CSF levels of the complement activation (cleaved) products [7,8,12–16] and thus our knowledge about CNS complement pathway activation is limited.

Semorinemab is a humanized IgG4 monoclonal antibody that binds to the N-terminal region of tau. The safety and efficacy of semorinemab has been evaluated in two Phase 2 studies, Tauriel (NCT03289143) and Lauriet (NCT03828747), in prodromal to mild (P2M) and mild to moderate (M2M) AD patients respectively [17,18]. In Tauriel, semorinemab did not slow clinical progression relative to placebo, as measured by the primary endpoint, the Clinical Dementia Rating sum of boxes score (CDR-SB) [17]. In Lauriet, the semorinemab arm had a significant reduction in cognitive decline measured by the Alzheimer’s Disease Assessment Scale-Cognitive Subscale 11 (ADAS-Cog11, co-primary endpoint) relative to the placebo. However, no treatment effect was observed on the functional outcome Alzheimer’s Disease Cooperative Study-Activities of Daily Living scale (ADCS-ADL; co-primary endpoint) or on any of the secondary endpoints. Although CSF tau indices were significantly reduced in both Tauriel and Lauriet, semorinemab did not prevent the accumulation of NFTs as measured by Genentech Tau Probe 1 ([^18^F]GTP1). The cognitive benefit observed in Lauriet, in the absence of a Tau PET effect, raises the hypothesis that semorinemab may alleviate cognitive dysfunction by reducing soluble toxic tau species [18]. Recent studies propose the presence of aberrant oligomeric tau species within synapses and their involvement in glia-mediated synapse engulfment in the AD brain [19,20]. We hypothesized that semorinemab may impact synapse loss through a modulation of complement pathway activity, which could be reflected by a change in CSF complement levels specifically in Lauriet (M2M) but not Tauriel (P2M) AD patients.

Here we examined intact (inactive) and cleaved (active) CSF complement proteins C4, C3 and Factor B (FB) in cognitively normal (CN) as well as P2M and M2M AD patients. We investigated the relationship between CSF complement protein levels and neurodegeneration/neuroinflammation biomarkers, tau PET, magnetic resonance imaging (MRI) and clinical outcomes in AD patients at baseline. Moreover, we assessed the pharmacodynamic effect of semorinemab on the change from baseline of CSF complement proteins in AD patients. We report elevated CSF C4a, C3a, Bb, C4, C3 and FB levels in AD patients. Intact and cleaved C4 displayed the greatest increase in AD vs CN CSF, consistent with increased classical pathway activity in AD CSF. Baseline CSF complement protein levels were correlated with neuro-axonal degeneration and glial activation biomarkers. Semorinemab had no treatment effect on CSF complement proteins relative to the placebo arm.

## 2. METHODS

### 2.1 Participants and sample collection

#### CN subjects

CN CSF samples (from subjects with MMSE scores of 28 or above) were procured from PrecisionMed (Carlsbad, CA). We confirmed the amyloid negative status by analysis of the CSF Ab42/40 ratio. Only subjects with CSF Ab42/40 ratios above 0.06 were included in this study (Figure S1, [21]). CSF was collected via lumbar puncture and any visibly blood contaminated samples were discarded prior to centrifugation for 10 minutes at 2400 rpm at room temperature. The supernatant was stored in 1 mL aliquots at -80°C. A subset of samples were frozen prior to aliquoting.

#### P2M and M2M AD subjects from the semorinemab Ph2 Tauriel and Lauriet trials

AD CSF samples were collected from the semorinemab Phase 2 studies Tauriel in prodromal to mild (P2M) AD; and Lauriet in mild to moderate (M2M) AD. Tauriel, conducted from October 2017 to July 2020, included 457 participants in a randomized double-blind 18-month course of intravenous infusions of 1,500, 4,500, or 8,100 mg of semorinemab (or placebo) every 2 weeks for the first 3 infusions, followed by infusions every 4 weeks [17]. The longitudinal analysis performed in this study only included the placebo and 4,500 mg semorinemab arms. CSF was collected at baseline/screening, and follow-up weeks 49 and 73. Lauriet, conducted from December 2018 to February 2020, included 272 participants in a randomized, placebo-controlled double-blind study of intravenous infusions of 4,500 mg of semorinemab (or placebo) every 2 weeks for the first 3 infusions, followed by every 4 weeks for 48 or 60 weeks [18]. CSF was collected at baseline/screening, and follow-up weeks 49 and 61. AD diagnosis was established based on confirmation of cognitive impairment by Clinical Dementia Rating Global Score (CDR-GS) of 0.5 or 1 for Tauriel and 1 or 2 for Lauriet, Mini-Mental State Examination (MMSE) clinical scores of 20 to 30 for Tauriel and 16 to 21 for Lauriet (inclusive), and amyloid pathology by PET scan or CSF Aβ(1-42) ≤1000 pg/mL by Elecsys® β-Amyloid (1-42) CSF immunoassay (Roche Diagnostics International Ltd, Rotkreuz, Switzerland) [17,18]. CSF was collected at the study sites via lumbar puncture directly into sterile, low-binding tubes. The first 1-2 mL, or more if necessary, of CSF was discarded until blood cleared from the CSF. After collection, CSF was mixed by gently inverting the tubes, and then centrifuged for 10 minutes at 2000 x g at 4°C. Supernatant was collected and aliquoted in 0.5 mL amounts, then stored at -70°C.

For the cross sectional analyses, baseline CSF samples from 79 Tauriel and 71 Lauriet participants were utilized. The longitudinal analyses included 39 (21 placebo, 18 semorinemab 4500 mg) and 47 (19 placebo, 28 semorinemab 4500 mg) participants from Tauriel and Lauriet, respectively. Note that only a subset of samples with both baseline and follow-up CSF were available at the time of this study, and thus the sample number included in the longitudinal analysis differs from Teng et al. 2022 [17] and Monteiro et al. 2023 [18]. Moreover, the number of available protein measurements per participant varies due to failed QC (see acceptance criteria in the assay sections below).

### 2.2 Aβ42/40 ratio measurements in CN CSF samples

CSF concentrations of Aβ(1-40) were measured using INNOTEST β-AMYLOID(1-40) (Cat. No. 81585; Fujirebio; Tokyo, Japan) at Genentech. Dilution linearity testing with CN CSF demonstrated %CV less than 15% for dilution factors of 1:100 to 1:200 measured in duplicate. Samples were diluted 1:100 and measured following the manufacturer’s protocol. Concentrations were interpolated from a 8-point calibration curve ranging from 7.8 to 1000 pg/mL. Acceptance criteria required less than 20%CV for duplicate sample measurements.

CSF concentrations of Aβ(1-42) were measured using INNOTEST β-AMYLOID(1- 42) (Cat. No. 81583; Fujirebio; Tokyo, Japan) at Genentech. Samples were measured neat following the manufacturer’s protocol. Concentrations were interpolated from a 6- point calibration curve ranging from 62.5 to 4599.2 pg/mL. Acceptance criteria required less than 20%CV for duplicate sample measurements.

### 2.3 pTau217 measurements in P2M and M2M CSF samples

CSF concentrations of phosphorylated tau (pTau217) were measured at Genentech using a custom single molecule array assay (Simoa, Quanterix Corp, Boston, MA). The reagents consisted of paramagnetic carboxylated beads coated with a rabbit monoclonal antibody specific for the phosphorylated T217 epitope of tau (Roche Diagnostics GmbH, Penzberg, Germany) and a biotinylated mouse monoclonal detection antibody specific for the mid-domain of tau (125B11H3, Genentech, Inc., South San Francisco, CA). Bead and antibody conjugations used the standard concentration and challenge ratios recommended by Quanterix, and the assays were run using a standard 2-step protocol on a Simoa HD-1 instrument (Quanterix). In the protocol, antibody-coated capture beads were incubated with diluted CSF (1:4) and the biotinylated detection antibody. After a wash step, the immunocomplexes were incubated with streptavidin-conjugated β- galactosidase (Quanterix), washed, re-suspended in resorufin β-D-galactopyranoside (Quanterix), and then applied to Simoa discs. The HD-1 analyzer was then used to read the resulting fluorescent signal and calculate the average number of enzymes per bead (AEB) for tested samples which were analyzed against an 8-point calibration curve constructed from GSK-3β-phosphorylated recombinant tau441 (SRP0689, SigmaAldrich, St. Louis, MO) ranging from 0.041 to 90 pg/mL. Samples were analyzed in duplicate using a single batch of reagents, with all available samples from a single participant measured in the same run. Qualification analyses demonstrated good detectability in human CSF (1.7-16.6 pg/mL, n=10 samples), inter-run precision (0.7-12%, n=10 samples, 2 runs), and parallelism (dilution linearity from 1:2 to 20, n=10 samples). The assay was not impacted by the presence of semorinemab (tested up to 6 mg/mL, which was approximately 1000 times the mean semorinemab trough concentration in CSF at weeks 49 and 73 following an 8100 mg dose). Acceptance criteria required less than 20%CV for duplicate sample measurements.

### 2.4 Elecsys and NeuroToolKit (NTK) measurements in P2M and M2M CSF samples

Concentrations of CSF Aβ42, Total-tau and Phosphorylated-tau (pT181) were measured blinded with the CE-marked Elecsys® β-Amyloid (1-42) CSF, Total-Tau CSF, and Phospho-Tau(181P) CSF in vitro diagnostic immunoassays on fully-automated cobase 601 analyzers (Roche Diagnostics International Ltd, Rotkreuz, Switzerland). The assays were validated to the Clinical and Laboratory Standards Institute (CLSI) Guidelines and exhibited a high level of within-run (<1.8%) and between-run (<2.3%) precision, which enabled singlicate measurements. Concentrations were interpolated from 2-point calibration curves, with measuring ranges of 200-1700 pg/mL (β-Amyloid (1-42)), 80.0- 1300 pg/mL (Total-Tau), and 8.00-120 pg/mL (Phospho-Tau(181P)). Acceptance criteria required two recombinant protein controls to quantify within 21% of their established concentrations. All measurements were performed in a blinded fashion at Covance CLS sites in Geneva (samples originating in Europe) and Indianapolis, IA (sample originating in the United States).

Concentrations of CSF Aβ40, α-synuclein, GFAP, IL-6, neurogranin, NfL, NPTX2, SNAP-25, S100b, sTREM2, and YKL-40 were measured using the NeuroToolKit robust prototype assays on fully-automated cobas e 411 and e 601 instruments (Roche Diagnostics International Ltd, Rotkreuz, Switzerland). The NeurotoolKit is a panel of exploratory prototype biomarker assays designed to specifically and robustly measure levels of soluble proteins associated with AD pathology, neuroinflammation, and neurodegeneration. The NeuroToolKit robust prototype assays are not validated to CLSI standards and are not CE-marked, but demonstrate high within-run (<5%) and between-run (<5%) precision, which also enabled singlicate measurements. Concentrations were interpolated from 5-point calibration curves, and acceptance criteria required two recombinant protein controls to quantify within 21% of their established concentrations. The NeuroToolKit measurements were performed in a blinded fashion at Microcoat Biotechnology (Bernried am Starnberger See, Germany) and Labcorp TBS (Greenfield, IN, USA).

#### Complement measurements in CN, P2M and M2M AD CSF samples

Because FT cycles affect complement protein concentrations (Gutierrez et al., under review), the number of FT cycles was matched for the CN and AD CSF. Previously developed and qualified ELISAs that measure C3, C3a, C4 and FB in CSF (Gutierrez et al., under review) were used for this study. For each ELISA, coating antibodies were applied to 96-well half-area high bind microplates (Corning), which were then incubated at temperatures between 2–8°C for durations ranging from 18 to 48 h. The plates were subsequently washed and blocked for a duration of 1 h. Next, diluted standards, controls, and samples were introduced into the microplate wells and were allowed to incubate at room temperature (RT) for a period of 1.5 h. Post-wash, HRP-conjugated detection antibodies (Peroxidase Labeling Kit - NH2, Dojindo, Cat. No. LK11) were incorporated into the C3, C3a, and C4 assays, and a biotinylated detection antibody (Biotin Labeling Kit - NH2, Dojindo, Cat. No. LK03) was incorporated into the FB assay. Following a 1 h incubation at RT the C3, C3a, and C4 assays underwent a final wash step. Meanwhile, a streptavidin poly-HRP solution was incorporated to the FB assay post-wash and allowed to incubate for an additional 30 min at RT. After the final series of washes, TMB substrate solution was added into the microplate wells to develop color, a process arrested by adding 1M phosphoric acid. The optical density of each well was calculated at 450 nm, utilizing a reference wavelength subtraction of 650 nm, using a SpectraMax ELISA plate reader (Molecular Devices). A logistic (4-PL) curve fit was applied to establish a standard regression curve. The concentrations of C4a were measured using the BD OptEIA Human C4a ELISA Kit (BD Biosciences, Cat. No. 550947), which utilizes a sandwich ELISA technique, as per the product guidelines. The concentrations of Bb were realized using a custom single molecule array Quanterix Simoa assay, as has been previously reported [22]. Each of the assays was subjected to a fit-for-purpose qualification and showed sensitivity to ex vivo complement activation (Gutierrez et al., under review). Samples with insufficient volumes and measurements with high %CV (>30%) or below the limit of quantitation were removed from analysis.

#### Tauriel/Lauriet clinical outcome measures used for correlation analyses

Cognitive/functional impairments were assessed at baseline and follow-up by ADCS-ADL (co-primary efficacy endpoint in Lauriet), ADAS-Cog13 (Tauriel only), ADAS-Cog11 (co-primary efficacy endpoint in Lauriet), MMSE and CDR-SB (primary efficacy endpoint in Tauriel) (see [17,18] for further details). The ADCS-ADL scale quantifies the performance of activities of daily living (ADL) in patients with AD and is administered to the care partners (score range 0–78; higher scores indicate better function) [23]. The ADAS- Cog13 and ADAS-Cog11 are 13-and 11-item cognitive scales, respectively, administered to assess cognitive domains most often affected in AD (score range 0-85, for ADAS- Cog13, and 0–70 for ADAS-Cog11, lower scores indicate better cognitive performance) [24]. The MMSE total score comprises subscores representing each cognitive domain: memory, orientation, attention, language, and construction (score range 0-30, lower scores indicate lower cognitive performance) [25]. The CDR–Sum of Boxes (CDR-SB) assesses six cognitive/functional domains (memory, orientation, judgment and problem solving, community affairs, home and hobbies, and personal care). Scores range from 0 to 18, with higher scores indicating greater impairment [26].

Magnetic resonance imaging (MRI) scans were performed at screening and follow-up, and included T1, T2*, and T2 fluid-attenuated inversion recovery sequences. Analyses were performed by a central MRI vendor (NeuroRx). T1 sequences were used to determine change from baseline to week 49, 61 or 73 in ventricular, whole brain, cerebral cortex, and hippocampal volumes (see [17,18] for further details). In this study, only whole brain measurements were used for the correlation analyses.

A subset of participants underwent [^18^F]GTP1 tau PET ([27]) at baseline and follow up at week 49 and 73 (for Tauriel), and at baseline and week 49 or 61 (for Lauriet) [17,18]. A central PET vendor (Invicro) determined [^18^F]GTP1 standardized uptake value ratio (SUVRs) across a whole cortical gray region of interest using cerebellar gray matter as reference (without partial volume correction).

### 2.5 Statistical analyses

Density distribution plots were generated to determine the distribution of CSF complement levels for each disease group. Log2 transformations were applied on single analyte measurements as skewed distributions were observed for raw values. The missing age of one subject from Tauriel (P2M AD) was imputed with the trial average age. For the cross sectional analyses, age and gender adjustment on reported CSF complement measurements were conducted by fitting linear regression models to the log2 transformed values. Residuals of the models were computed and used to calculate a constant equal to the mean of the log2 transformed value for each protein minus the mean of the residuals. The residuals were adjusted by adding the calculated constant values and these adjusted residuals were used in the P2M and M2M AD vs. CN comparisons (Figure 2, Figure S4). Two tailed Welch’s t-tests were used to compare mean differences between groups (Figure 2, Figure S2, Figure S5, Figure 4, Figure 5) with α = 0.05. Boxes represent the median and interquartile range (IQR); the lower and upper hinges correspond to the first and third quartiles (the 25th and 75th percentiles). The upper whisker extends from the hinge to the largest value no further than 1.5 * IQR from the hinge. The lower whisker extends from the hinge to the smallest value at most 1.5 * IQR of the hinge. Individual data points are plotted over the box plots. Spearman’s rank correlation coefficient ρ was calculated for all correlation analyses with α = 0.05. When applicable, adjustment for multiple testing was completed via the FDR method.

**Figure 1:**
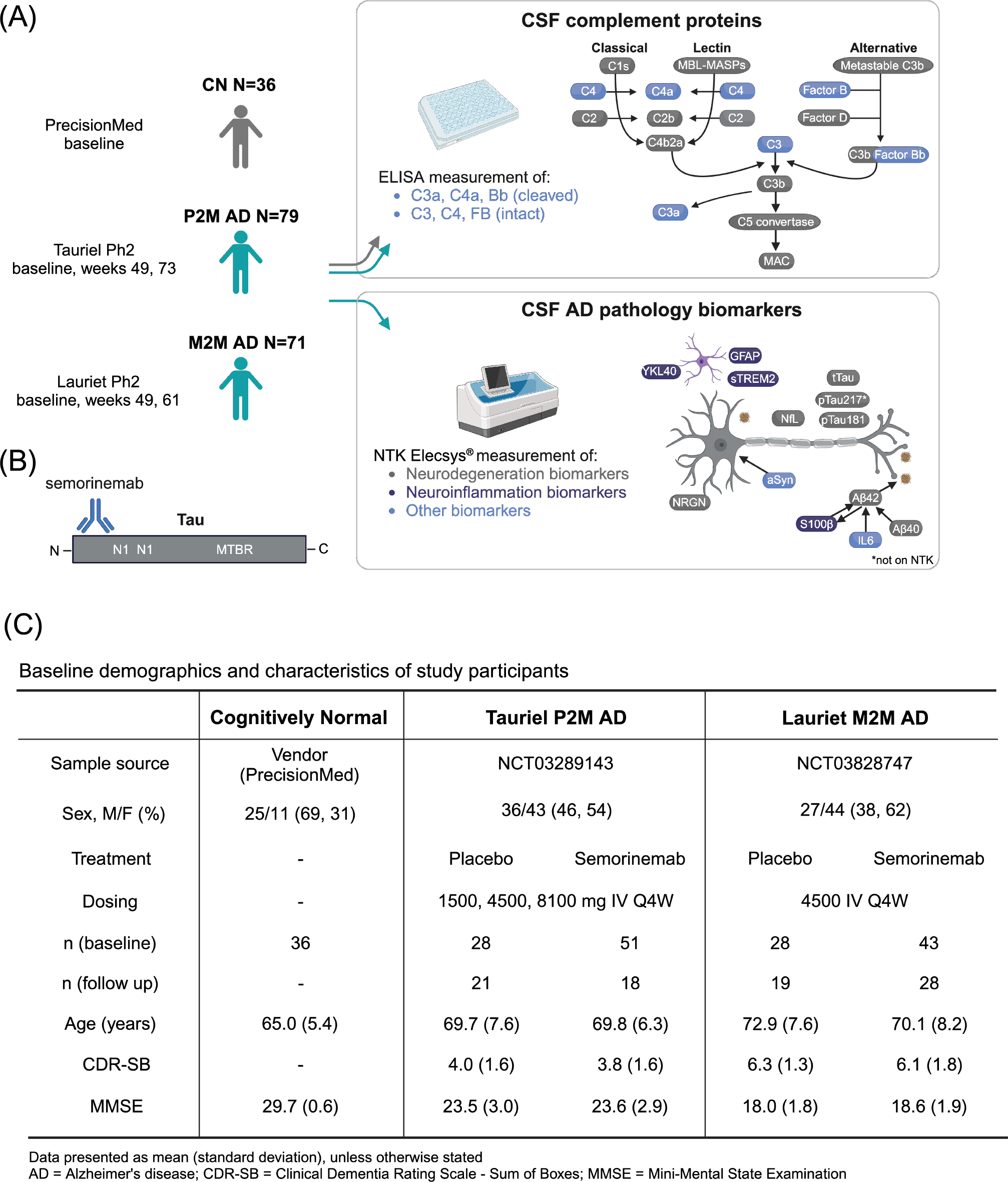
**(A)** CSF complement levels from cognitively normal (CN, n=36), prodromal to mild AD (P2M, Tauriel, n=79), and mild to moderate AD (M2M, Lauriet, n=71) participants were measured by immunoassay. CSF neurodegeneration and neuroinflammation biomarkers were measured for the two AD groups using the Roche NeuroToolKit (NTK), Elecsys in vitro diagnostic immunoassays (Aβ42, tTau, pTau181), and ELISA (pTau217). (**B)** Semorinemab is an IgG4 antibody binding to the N-terminal region (residues 6-23) of all six isoforms of human tau, including phosphorylated species. (**C)** Baseline demographics and characteristics of the CN, P2M AD and M2M AD subjects included in this study. This figure was created with BioRender.com.

**Figure 2:**
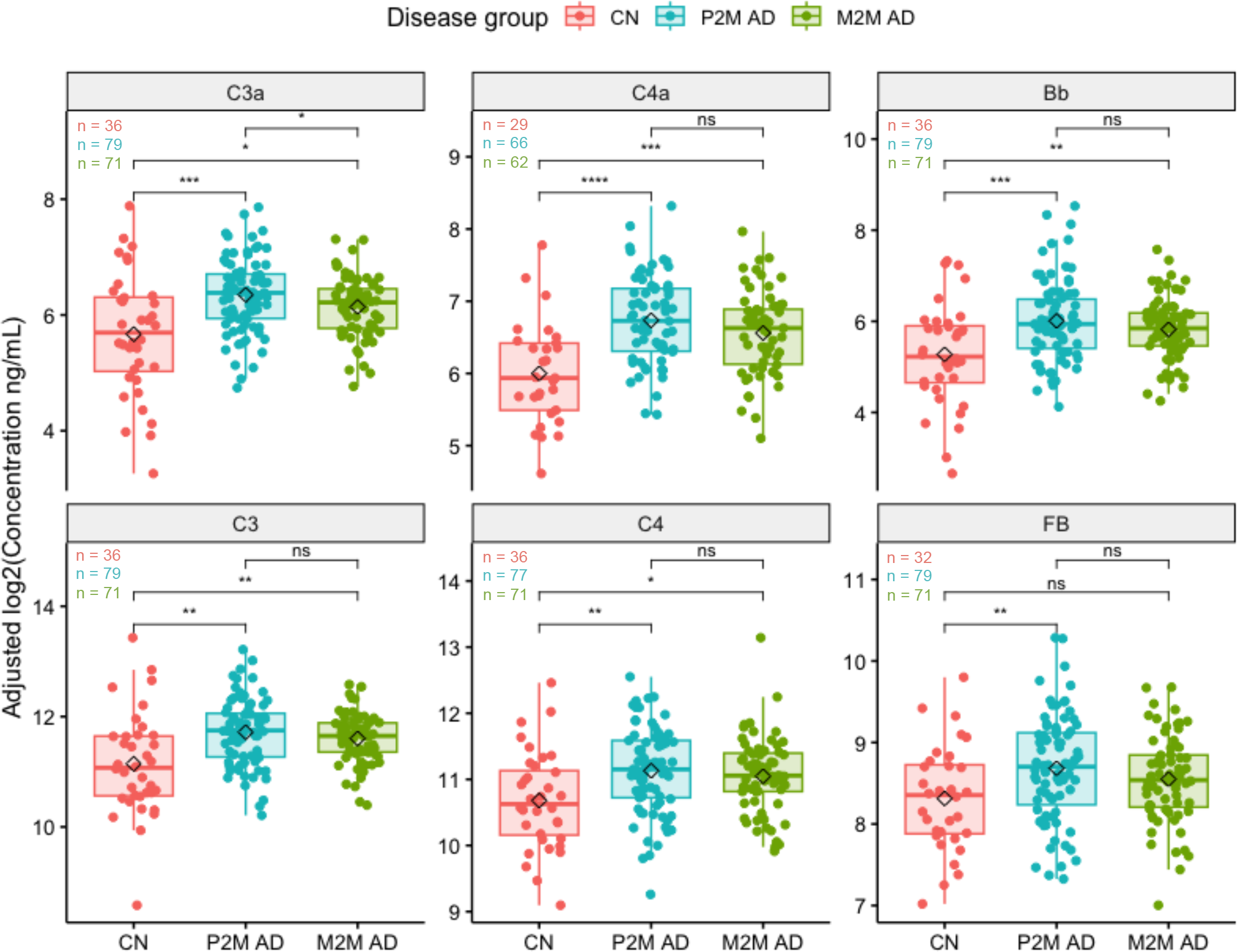
Box plots of age and gender adjusted baseline CSF complement concentrations log2(ng/mL) from P2M AD, M2M AD, and CN participants. Black diamonds represent mean values of each disease group. *p<0.05, **p<0.01, ***p<0.001, ****p<0.0001, ns (not significant) p ≥ 0.05, t-test. See table S1 for absolute concentrations.

Complement protein measurements from predose CSF from both the placebo and semorinemab arms were included in the correlation analysis of baseline complement proteins with baseline NTK proteins, vMRI, ^18^F]GTP1 tau PET SUVRs or cognitive/functional clinical scores (including CDR-SB, ADAS-Cog, MMSE and ADCS-ADL) (Figure 3B and C). The baseline correlation analyses between complement protein levels and ADAS-Cog13 or ADAS-Cog11 were represented by trial as Tauriel and Lauriet each only measured one of these scores (ADAS-Cog13 for Tauriel, ADAS-Cog11 for Lauriet) (Figure 3B). In the correlation analysis of the changes from baseline at follow-up in complement proteins with the changes from baseline at follow-up in NTK proteins, semorinemab-treated patients were not included as treatment effects were observed longitudinally on the NTK proteins for both studies (Figure S6A). Due to the low N of placebo-treated participants with both baseline and follow up measurements, semorinemab-treated participants from Tauriel were included in the correlation analysis of changes from baseline at follow-up of the complement proteins with the changes from baseline at follow-up in vMRI, ^18^F]GTP1 tau PET SUVRs or cognitive/functional clinical scores (including CDR-SB, ADAS-Cog, MMSE and ADCS-ADL) (Figure S6B). Because no treatment effect was observed, we considered this approach reasonable. Semorinemab-treated participants from Lauriet were excluded because of the treatment effect observed in ADAS-Cog11. Unlike the baseline analysis, the correlation analysis with the change from baseline in ADAS-Cog could not be grouped by trial due to the low longitudinal sample count so the percent change from baseline was used instead to account for the difference in score ranges. For correlation analyses of change from baseline at follow-up in complement proteins with changes from baseline at follow-up in NTK proteins and changes in cognitive/functional scores (except ADAS-Cog), absolute changes were used (Figure S6A and S6B).

**Figure 3:**
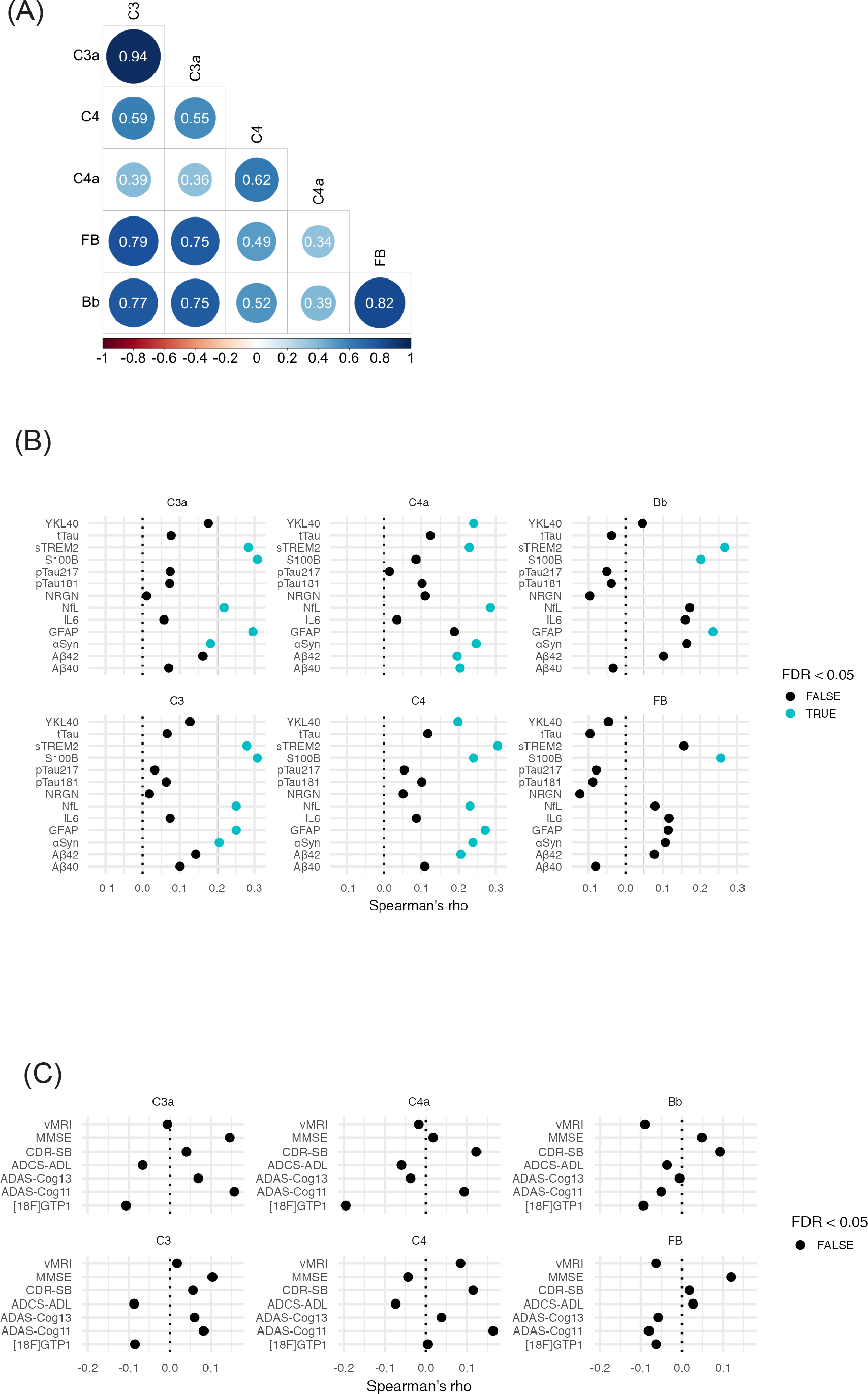
**(A)** Heatmap of the correlation between CSF complement levels in baseline P2M AD and M2M AD participants. Spearman’s rank correlation coefficient values are displayed and circles represent significant correlations with FDR-adjusted p-values < 0.05. **(B)** Spearman’s rank correlation between CSF complement and NTK proteins in P2M and M2M AD participants at baseline. Blue and black dots indicate the FDR adjusted p-value less than or greater than 0.05, respectively. **(C)** Spearman’s rank correlation between CSF complement and cognitive/functional scores, [18F]GTP1 tau-PET SUVRs, and whole brain vMRI scan measurements in P2M and M2M AD participants at baseline. Adas-Cog13 and Adas-Cog11 were used in Tauriel (P2M AD) and Lauriet (M2M AD), respectively [17,18]. Blue and black dots indicate the FDR adjusted p-value less than or greater than 0.05, respectively.

**Figure 4:**
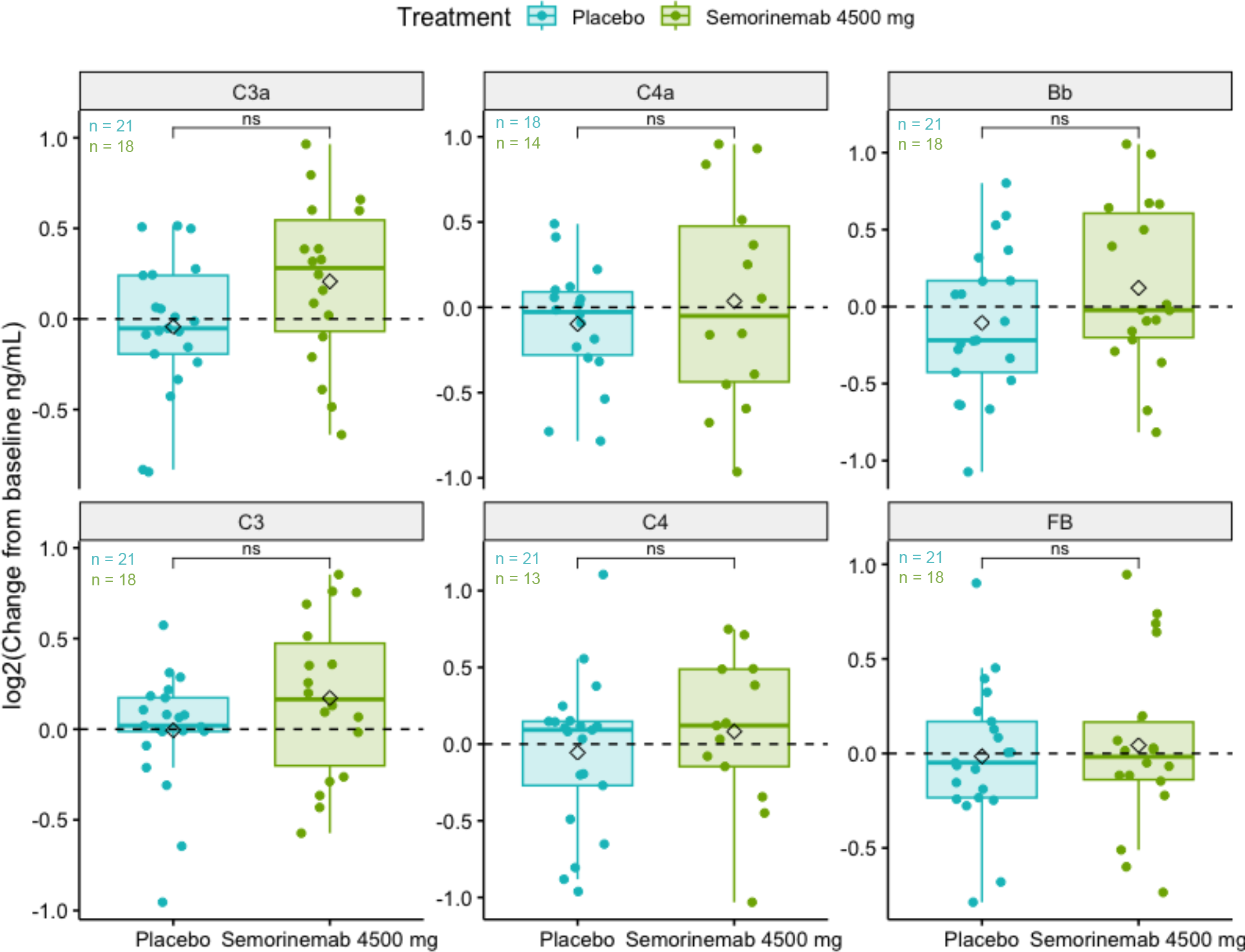
Box plots of log2(change from baseline at weeks 49 or 73 in CSF complement concentrations ng/mL) of Tauriel P2M AD patients treated with placebo or semorinemab at 4500 mg. Black diamonds represent mean values for each treatment per analyte. *p<0.05, **p<0.01, ***p<0.001, ****p<0.0001, ns (not significant) p ≥ 0.05, t-test.

**Figure 5:**
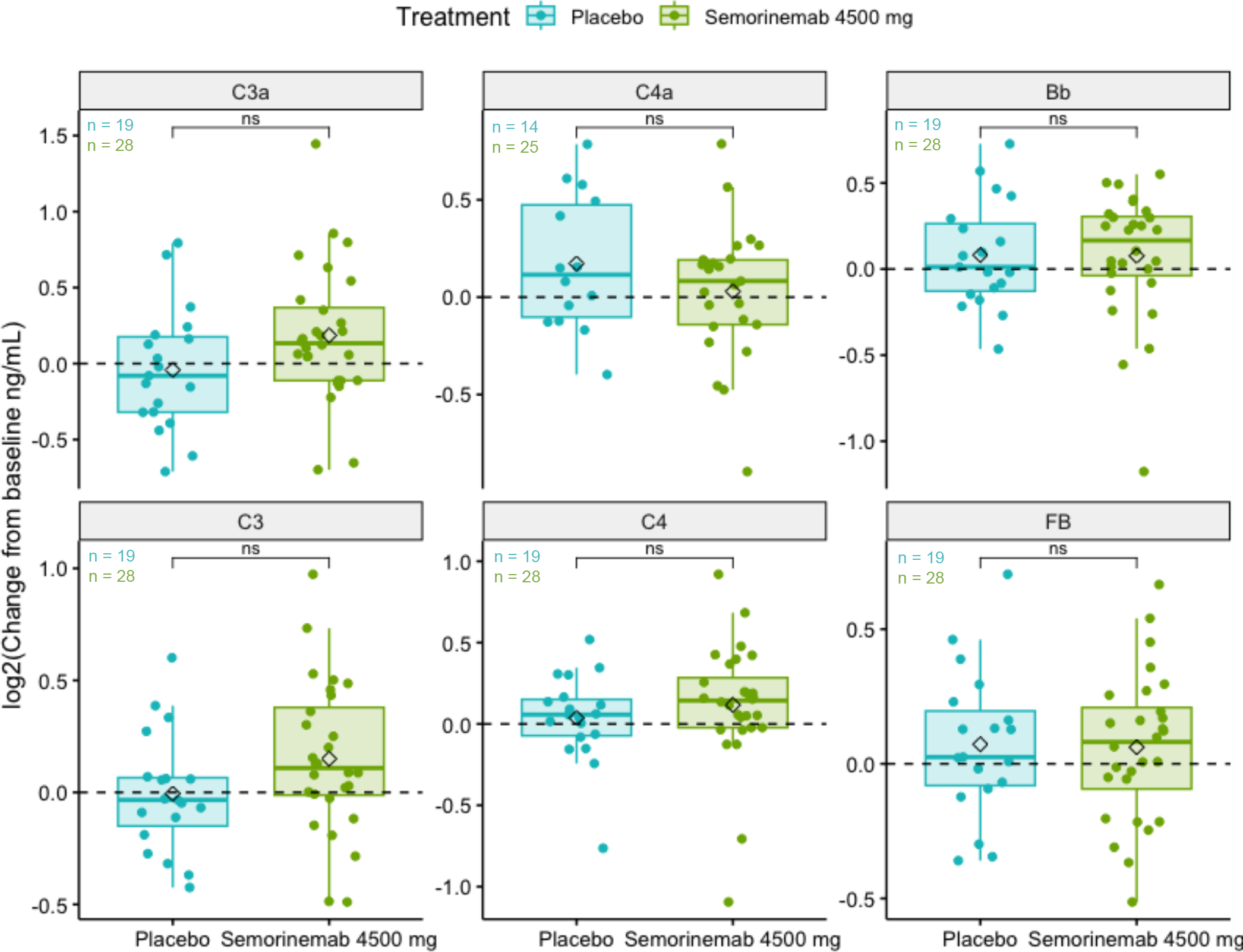
Box plots of log2(change from baseline at weeks 49 or 61 in CSF complement concentrations ng/mL) of Lauriet M2M AD patients treated with placebo or semorinemab at 4500 mg. Black diamonds represent mean values for each treatment per analyte. *p<0.05, **p<0.01, ***p<0.001, ****p<0.0001, ns (not significant) p ≥ 0.05, t-test.

All analyses were completed using publicly available packages in Python version 3.11.4 and R version 4.3.1.

## 3. RESULTS

### Study design, participant characteristics and CSF analytes

The present study included CSF samples from CN, P2M AD (Tauriel) and M2M AD cohorts (Lauriet) (Figure 1A). For the CN cohort, CSF was collected at a single time-point. Tauriel/Lauriet participant CSF was collected at baseline and following treatment with placebo or semorinemab (Figure 1A and B). Immunoassays were used to measure the levels of intact (inactive) and cleaved (active) complement proteins C4, C3 and FB (Figure 1A). Neurodegeneration and neuroinflammation biomarkers were evaluated using the NTK panel or by immunoassay ([28], Figure 1A).

The CN cohort included 69% males, while the P2M and M2M AD cohort included 46% and 38% males, respectively (Figure 1C). Despite attempts to age-match the CN and AD cohorts, the CN participants were on average 4.7-7.9 years younger than the AD patients (Figure 1C). Because previous studies showed that peripheral complement protein levels were influenced by age and gender [29], we first sought to investigate the impact of these demographic characteristics on complement levels. CSF complement protein levels C3a, C3, Bb and FB were mildly elevated in male subjects relative to females (Figure S2) with median fold differences of 1.17, 1.20, 1.42, and 1.22, respectively (Table S1). CSF C4a, C4, C3a, C3 and Bb measurements showed a weak (Spearman Rho values ranging between 0.18 and 0.35) but significant correlation with age (Figure S3).

### Baseline CSF complement measurements in CN and AD subjects

We next assessed the differences in age/gender adjusted CSF complement protein levels (see Methods) between CN and baseline P2M or M2M AD subjects. Baseline CSF complement proteins C4a, C4, C3a, C3 and Bb levels were increased in P2M and M2M AD patients relative to CN subjects (Figure 2, Figure S4, Table S1). Among these analytes, C4a levels displayed the largest fold-increase between CN and AD CSF, with a 2.2-fold increase in P2M AD CSF, and a 1.9-fold increase in M2M AD CSF (Figure 2, Table S1). CSF FB showed a mild but significant elevation in P2M but not M2M AD patient samples (Figure 2, Table S1). None of the tested complement proteins significantly differed between P2M and M2M AD patients. These data suggest that C4, C3 and FB expression and activation are higher in the CSF of AD patients (vs CN), with the greatest difference observed for C4a.

### Baseline CSF complement proteins and neurodegeneration/neuroinflammation biomarkers in AD patients

Next, we investigated the intercorrelation patterns for different complement proteins as well as their association with neurodegeneration and neuroinflammation biomarkers [30] in the CSF from P2M and M2M AD patients. As expected, the highest correlations were found between the intact and cleaved forms of each complement protein (Spearman Rho > 0.6) (Figure 3A). Moderate to strong associations were also found between different complement proteins: CSF C3a and C3 displayed the strongest correlation with Bb and FB (Figure 3A), likely due to the involvement of cleaved C3 in the alternative pathway activation (Figure 1A). CSF C4a and C4 were found to be more moderately associated with C3a, C3 and FB (Figure 3A).

When examining the correlations between complement proteins and neurodegeneration/neuroinflammation biomarkers, C4, C3a and C3 were found to be modestly associated with NfL (Neurofilament Light), GFAP (Glial Fibrillary Acidic Protein), ɑSyn (alpha-Synuclein), sTREM2 (soluble Triggering Receptor Expressed on Myeloid cells 2) and S100b (S100 calcium-binding protein B) (Figure 3B, Spearman Rho = 0.21- 0.31). C4a was moderately correlated with Aβ40, Aβ42, ɑSyn, NfL, sTREM2 and YKL40 (Chitinase-3-like protein) (Figure 3B, Spearman Rho = 0.2 - 0.29). CSF Bb levels were associated with GFAP, sTREM2 and S100b levels (Spearman Rho = 0.2-0.27), while FB only correlated with S100b (Spearman Rho = 0.26) (Figure 3B). No significant correlations were observed between the tested complement proteins and tau indices (tTau, pTau181 and pTau217).

### Baseline CSF complement proteins and MRI, tau PET and clinical outcome measures in AD patients

None of the CSF complement proteins showed a meaningful correlation with whole brain volumetric MRI (vMRI) measurements, [^18^F]GTP1 tau PET SUVRs or cognitive/functional clinical scores (including CDR-SB, ADAS-Cog (ADAS-Cog13 for Tauriel and ADAS- Cog11 for Lauriet), MMSE and ADCS-ADL) at baseline (Figure 3C). Together, our findings indicate that baseline CSF complement proteins correlate with baseline CSF neurodegeneration/neuroinflammation biomarker levels but not with brain atrophy or cognitive/functional scores.

### Longitudinal changes in CSF complement levels in placebo and semorinemab- treated AD patients

In the placebo arms from Tauriel (P2M) and Lauriet (M2M), CSF C4a, C3a, Bb, C4, C3 and FB levels did not significantly change from baseline to follow-up (weeks 49, 61 or 73) (Figure S5A and B). No consistent meaningful correlation patterns were found between the changes from baseline in complement proteins and changes from baseline in neurodegeneration or neuroinflammation biomarkers (Figure S6A), [^18^F]GTP1 tau PET SUVRs, whole brain vMRI, functional or clinical scores (Figure S6B). The change from baseline of CSF complement proteins C4a, C3a, Bb, C4, C3 and FB did not significantly differ between semorinemab at 4500 mg and placebo in Tauriel P2M (Figure 4) or Lauriet M2M (Figure 5) AD patients. These data suggest that complement proteins do not significantly change from baseline at weeks 49, 61 or 73. Furthermore, we show that, despite the cognitive benefit observed in Lauriet, semorinemab does not modulate CNS complement activity in P2M (Tauriel) or M2M (Lauriet) AD patients.

## 4. DISCUSSION

The present study demonstrated elevated CSF complement protein levels in P2M/M2M AD when compared to CN, with the greatest difference observed for C4a. In AD patients, several CSF complement proteins were correlated with glial activation and neurodegeneration markers including sTREM2, GFAP, YKL40 and NfL. Despite showing a reduction in soluble tau in CSF, semorinemab did not modulate complement proteins relative to placebo. Our results provide evidence for increased complement pathway activity in AD patients and could aid the development of therapeutic interventions targeting the CNS immune system.

Among all tested complement proteins, CSF C4a showed the most robust difference, with higher levels observed in AD patients compared to CN subjects. We also observed significant increases of CSF intact C4, suggesting increased CNS C4 expression *and* activation in AD patients. Elevated CSF C4 levels are consistent with previous reports of increased expression of complement pathway components in glia from AD patients and mouse models [31,32]. C4 processing occurs downstream of the classical and lectin pathways [33]. While the lectin and alternative pathways are thought to contribute little to AD pathogenesis, numerous studies have reported increased levels of classical pathway activator proteins in the CNS from AD patients as well as amyloid/tauopathy mouse models [6–8,34]. Furthermore, the deletion or inhibition of the classical pathway initiator molecule C1q rescues microglia- and astrocyte-mediated synapse phagocytosis in mouse models of amyloidosis and tauopathy [7–9]. Taken together, these data suggest that the increases in CSF C4 and C4a (and C3a) could reflect enhanced expression and activity of classical complement pathway components in AD.

Contrary to previous reports showing an association between CSF or brain C3 levels and tau [7,35], we did not find a correlation between baseline CSF complement proteins and tau (both soluble CSF tau and aggregated tau measured by Tau PET) in AD patients. The inconsistency between our results and previous reports could be explained by differences in the number of study participants, the participant disease stage or the analytical methods. Notably, baseline CSF C4a, C3a, C4 and C3 correlated positively with the neurodegeneration biomarker NfL, consistent with recent findings in familial FTD [36]. Baseline AD CSF C4a, C3a, C4 and C3 also displayed a positive association with multiple glial activation biomarkers, including sTREM2, YKL40 and GFAP. Since brain complement pathway components are expressed and secreted by microglia and astrocytes [37], these data support the hypothesis that changes to CSF complement protein levels and activity reflect CNS neuroinflammation in AD.

CSF complement levels did not change over 49, 61 or 73 weeks in AD patients and semorinemab demonstrated no pharmacodynamic effect on CSF complement protein changes from baseline at week 49 when compared to placebo. Thus, despite evidence of target modulation in the CNS, there is no semorinemab-mediated impact on CNS complement activity. This finding suggests that the significant reduction in cognitive decline measured by ADAS-Cog11 in Lauriet may not be attributable to modulation of CNS complement activity.

Although our findings provide evidence for increased complement protein levels and/or activity in AD CSF, the present study has limitations. First, even though precautions were taken to align the preanalytical conditions for all groups as much as possible, the CSF collection and handling of the CN and Tauriel/Lauriet samples differed slightly. Second, the correlation analyses between CSF complement proteins and clinical outcomes were performed on a relatively small subset of AD subjects (Table S2). Third, there is a gender imbalance between the CN and AD groups, with greater numbers of male subjects in the CN and females in the AD cohorts. Since we reported mildly elevated CSF C3a, C3, Bb and FB levels in male participants, the observed AD-associated increases in these proteins may be underestimated due to the greater number of males included in the CN vs AD cohorts. Together these limitations warrant further studies to assess CSF C4a, C3a, Bb, C3, C4 and FB in larger cohorts with equal biofluid preanalytical handling and accessible clinical/biomarker from age- and gender-matched CN and AD subjects.

Overall, our results provide in vivo evidence for increased complement pathway activity in AD patient CSF. Future longitudinal studies including a presymptomatic patient population will be essential to determine at which disease stage the CNS complement pathway activity starts to increase in AD patients. Moreover, it will be important to elucidate how the CSF intact/cleaved complement protein levels relate to synapse loss, as determined by SV2A-PET.

## Data Availability

All data produced in the present study are available upon reasonable request to the authors

## Acknowledgements/Conflicts/Funding Sources

We thank Dan Abramzon, Rod Mathews, Jeannette Lo and Bill O’Gorman for comments on the manuscript.

CS, JL, JG, FY and AB: Study design. CS, ET, CM, JL, JG, RN, SS, GK: Sample and data acquisition. JL, BT and AB: Data analysis. CS, JL, BT, JG, JH, CM, ET, FY and AB: Data interpretation. CS, JL and AB: Manuscript writing - original draft. All authors: Manuscript writing - review and editing.

The NeuroToolKit is a panel of exploratory prototype assays designed to robustly evaluate biomarkers associated with key pathologic events characteristic of AD and other neurological disorders, used for research purposes only and not approved for clinical use (Roche Diagnostics International Ltd, Rotkreuz, Switzerland). Elecsys β-Amyloid (1-42), Total-Tau, and Phospho-Tau (181P) CSF assays are approved for clinical use.

COBAS and ELECSYS are trademarks of Roche. All other product names and trademarks are the property of their respective owners.

GK is a full-time employee of Roche Diagnostics GmbH, Penzberg, Germany. All other authors are full-time employees of Genentech Inc. (member of the Roche group). Edmond Teng is listed as a co-inventor on the patent for semorinemab.

---

**Figure S1:**
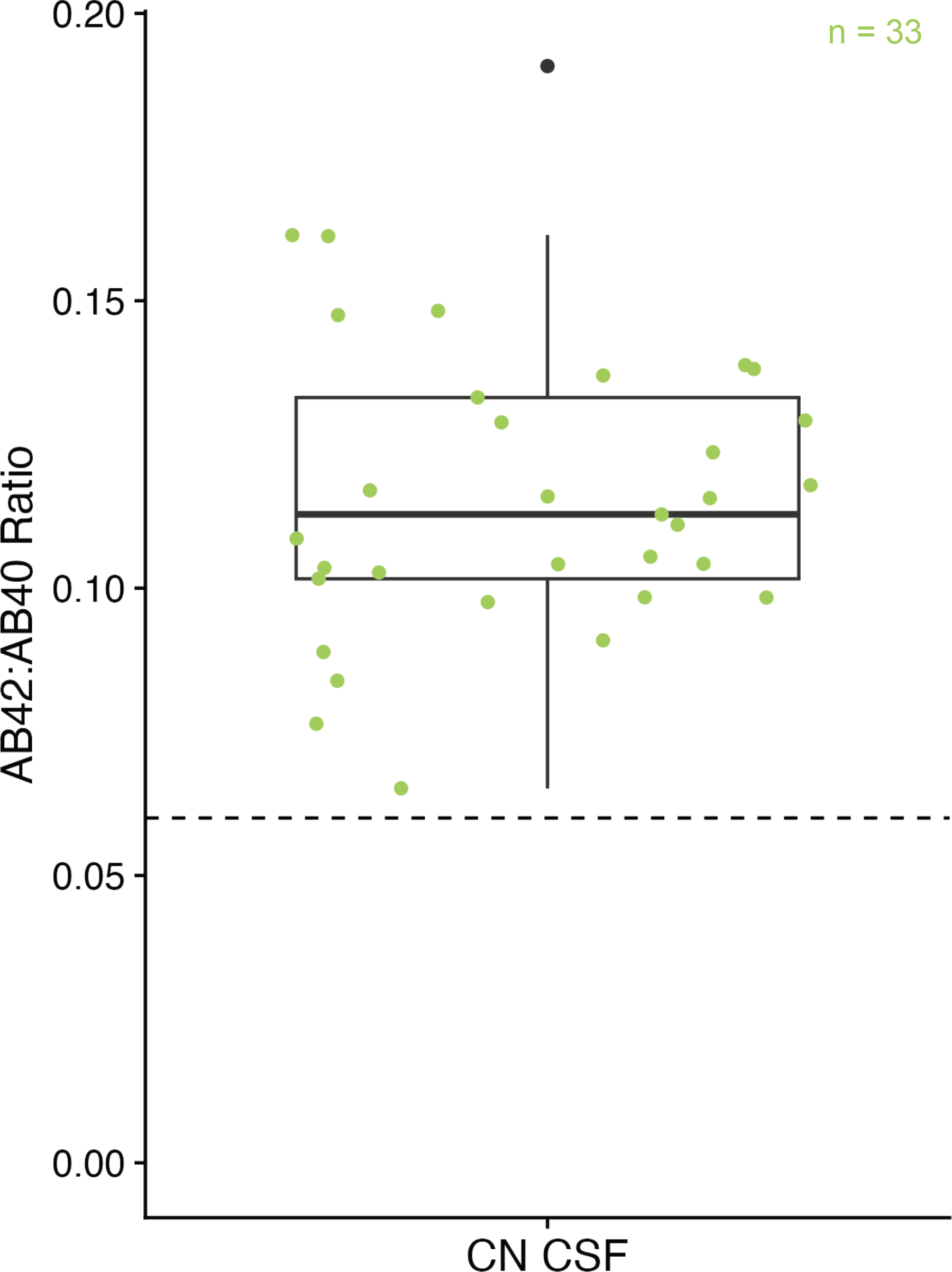
Box plot of CSF Aβ(1–42):Aβ(1–40) ratio of procured CN CSF (n=36). Dashed line refers to 0.06 cutoff value distinguishing between Aβ-positive (< 0.06) from Aβ- negative (>0.06) individuals.

**Figure S2:**
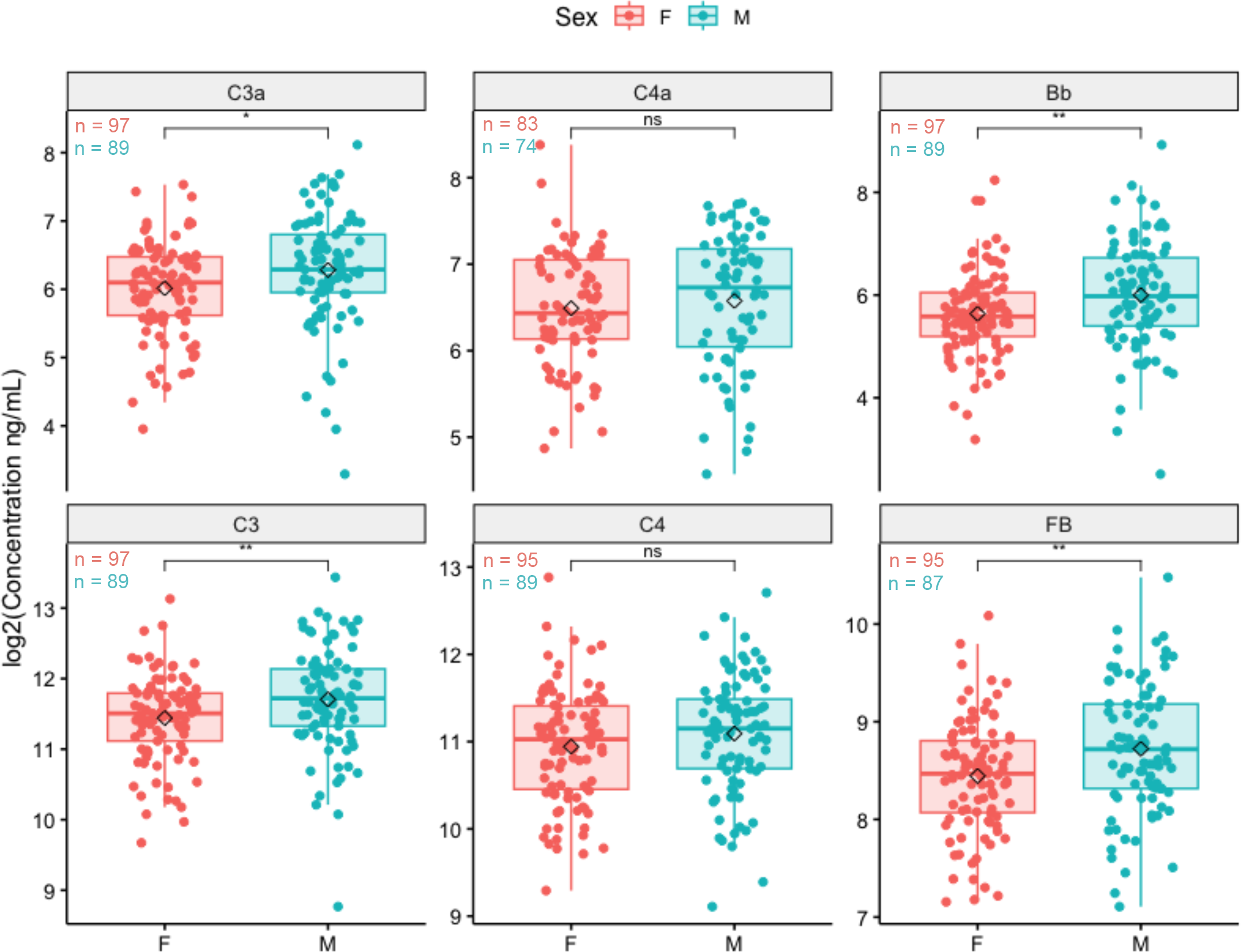
Box plots of baseline CSF complement concentrations log2(ng/mL) from P2M AD, M2M AD, and CN participants grouped by sex. Black diamonds represent mean values for males and females per analyte. *p<0.05, **p<0.01, ***p<0.001, ****p<0.0001, ns (not significant) p ≥ 0.05, t-test. See table S1 for absolute concentrations.

**Figure S3:**
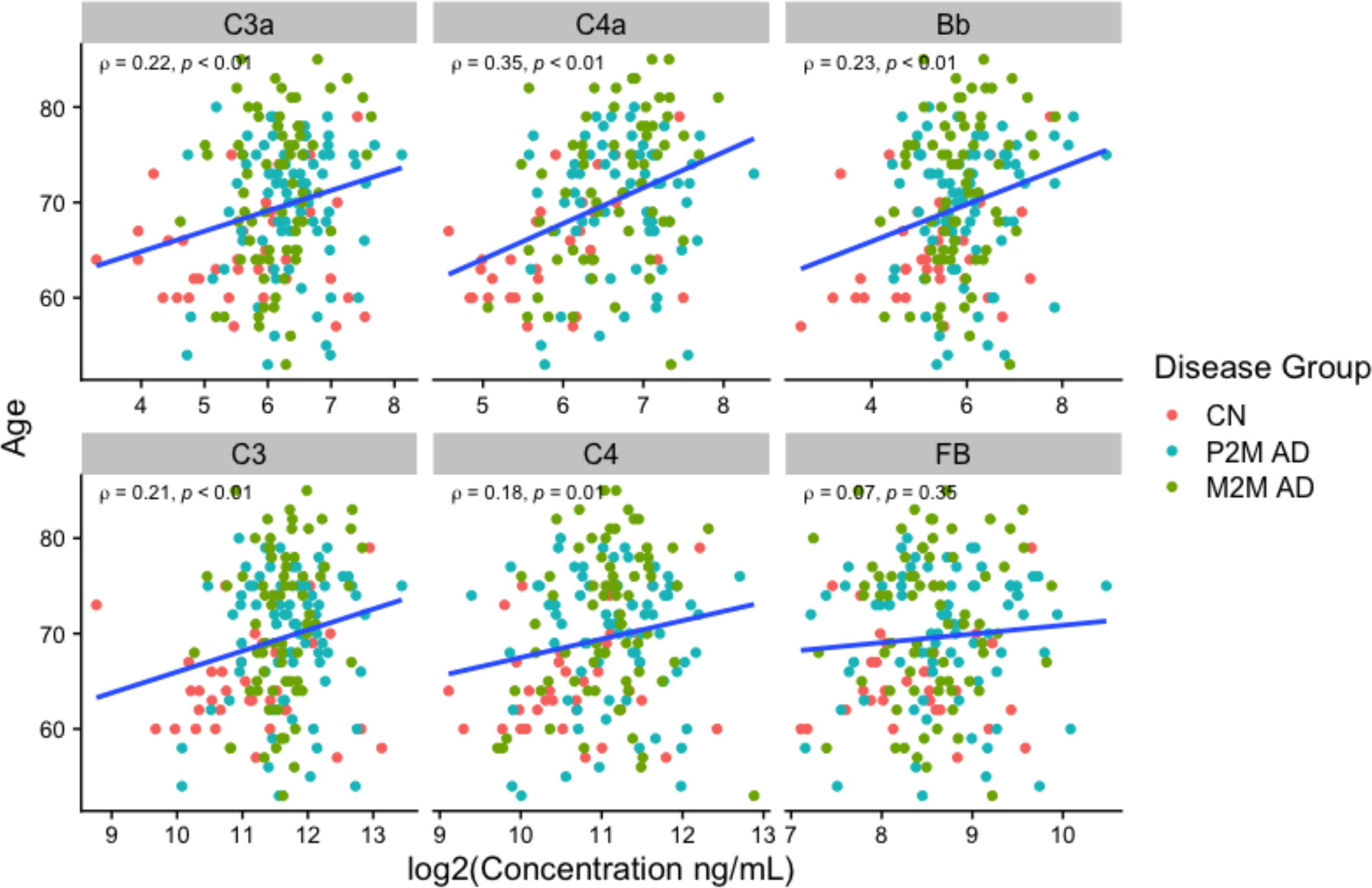
Spearman’s rank correlation between baseline CSF complement concentrations log2(ng/mL) and age of P2M AD, M2M AD, and CN participants. Blue lines represent the linear regression lines fitted between complement levels and age.

**Figure S4:**
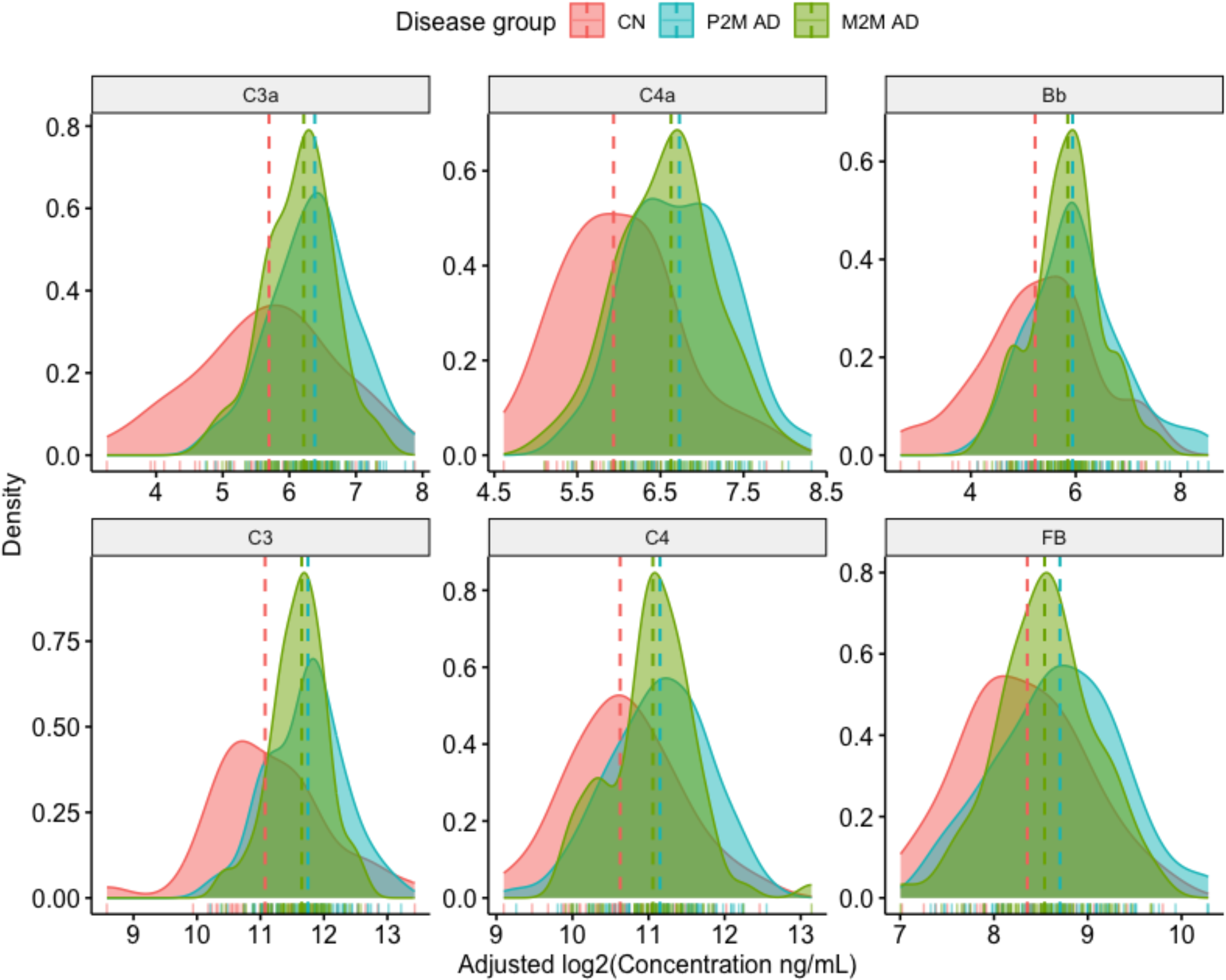
Density distribution of age and gender adjusted CSF complement levels by disease group. Dashed lines indicate the median value for each disease group.

**Figure S5:**
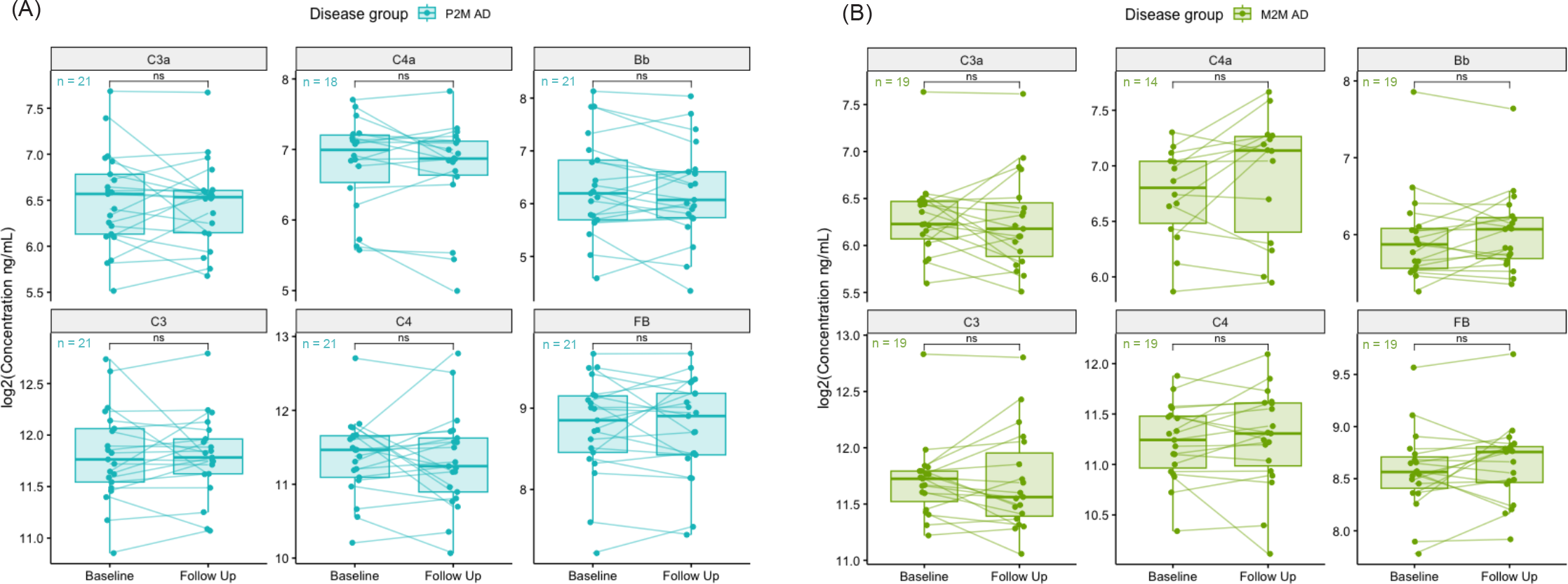
Box plots of CSF complement concentrations log2(ng/mL) of **(A)** Tauriel P2M AD patients and **(B)** Lauriet M2M AD patients treated with placebo at baseline and follow-up (weeks 49, 61 or 73). *p<0.05, **p<0.01, ***p<0.001, ****p<0.0001, ns (not significant) p ≥ 0.05, paired t-test.

**Figure S6:**
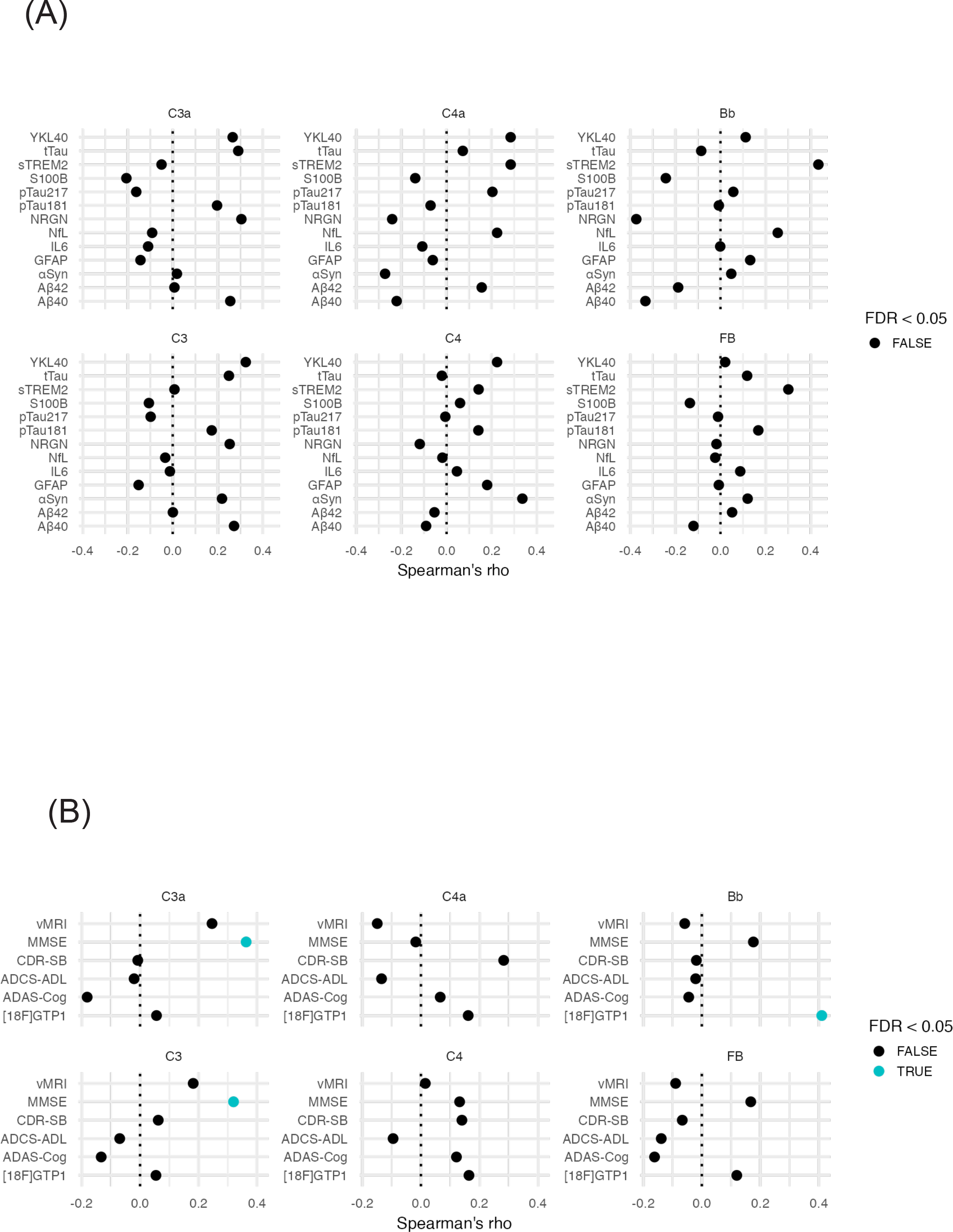
**(A)** Spearman’s rank correlation between the change from baseline at follow-up of CSF complement and the change from baseline at follow-up of NeuroToolKit proteins in P2M AD and M2M AD (placebo only). Blue and black dots indicate the FDR adjusted p-value less than or greater than 0.05, respectively. **(B)** Spearman’s rank correlation between the absolute change from baseline at follow-up in CSF complement levels and the change from baseline at follow-up in cognitive/functional scores, [18F]GTP1 tau-PET SUVRs, and whole brain vMRI scan measurements in P2M AD (Tauriel, placebo and semorinemab arms, see Methods) and M2M AD (Lauriet, placebo only, see Methods). Percent changes from baseline are represented for ADAS-Cog only as Adas-Cog13 and Adas-Cog11 were used in Tauriel and Lauriet, respectively [17,18]. Blue and black dots indicate the FDR adjusted p-value less than or greater than 0.05, respectively.

Table S1: Baseline absolute CSF complement concentrations (ng/mL) in CN, P2M AD, and M2M AD subjects and median CSF complement fold-changes between disease group or sex with the total count of non-missing values. Anonymized samples were received with unique subject identification numbers, and then randomized for analysis.

Table S2: Number of AD patients included in the correlation analyses from Figure 3 and Figure S6.

